# Multimodal hypersensitivity across the menarcheal transition is associated with pelvic pain in adolescents: a longitudinal study

**DOI:** 10.64898/2026.09.09.26362449

**Authors:** Matthew J. Kmiecik, Kevin M. Hellman, Sarah E. Darnell, Gregory E. Miller, Frank F. Tu

## Abstract

Multimodal hypersensitivity (MMH), heightened sensitivity across multiple sensory modalities, is associated with chronic pain risk in adults. How MMH develops across adolescence and whether early pain experiences shape its trajectory remains unknown. The Early Menstrual Pain Impact on Multisensory Hypersensitivity (EMPATHY) study followed 300 premenarcheal adolescents (age *M*=11.29, *SD*=0.98 years) across the menarcheal transition with three visits comprising a comprehensive multimodal sensory testing battery: visual and auditory unpleasantness, a non-invasive bladder filling task, pressure pain thresholds, after-pain ratings, cold pain, and conditioned pain modulation. A principal component (PC) analysis of 24 baseline measures (*n*=214) identified three components accounting for 34.2% of variance: MMH (PC1; 16.5% of variance), a pressure-pain stimulus-response function (PC2; 9.3%), and bladder hypersensitivity (PC3; 8.4%). All experimental measures, and four self-report sensory sensitivity questionnaires projected as supplementary variables, loaded onto PC1, supporting its interpretation as MMH and replicating the component structure previously observed in adult women. Longitudinal trajectories of MMH (*n*=140) were modeled with linear mixed-effects regression with menstrual pain, pelvic pain, time, and their interactions as predictors, adjusting for pubertal staging. MMH decreased across the menarcheal transition (*p*=0.029). Pelvic pain (*p*=1.24x10^-6^), but not menstrual pain (*p*=0.205), was positively associated with MMH, independent of time and pubertal development. These findings establish MMH as a measurable, replicable sensory phenotype in early adolescence that covaries with non-menstrual pelvic pain, and position it as a candidate early marker of nociplastic pain risk. Future studies are needed to support interventions and screening for elevated sensory sensitivity before pain becomes clinically entrenched.

---

Chronic pain affects 1 in 5 children and adolescents worldwide [11]. Pain during adolescence increases the risk of chronic pain in adulthood [26,71], suggesting that pain management and prevention should begin during transitional periods such as from middle-childhood to adolescence [51]. During this transitional period, puberty occurs and is marked by rapid changes in gonadal hormones thought to modulate pain sensitivity [38,48]. However, critical questions remain: How does pain system sensitivity develop during the pubertal transition? And what early pain experiences, such as the onset of menstrual pain, contribute to individual differences in that trajectory? Multimodal hypersensitivity (MMH) is one such measure that may capture individual differences in pain system sensitivity across development [15,33,50]. Unlike single-modality QST measures, MMH captures shared variance across sensory systems and may therefore index a more stable, trait-like dimension of pain system function.

MMH is increased sensitivity across multiple sensory modalities (e.g., vision, touch, hearing, and visceral sensation) and is associated with chronic pain conditions [31] and sensory modulation disorder [4]. MMH is a main component of nociplastic pain [18] and is thought to reflect the neurobiological underpinnings of central sensitization [37]. Unlike subjective patient-reported outcomes used to assess generalized sensory sensitivity (GSS) [59], MMH objectively quantifies cross-modality hypersensitivity using experimental pain measurements. In adult females, MMH was associated with future pelvic pain and patient-reported outcomes of genitourinary symptoms, depression, anxiety, and health [33].

Moderate to severe menstrual cramping pain, dysmenorrhea, is a general risk factor for chronic pain [40] with severe dysmenorrhea in adolescence associated with a 76% increased risk in chronic pain at age 26 relative to those reporting no dysmenorrhea [55]. Dysmenorrhea was associated with MMH and GSS in a cohort of adults with co-morbid pain conditions [33,58]. In adolescents, postmenarcheal widespread bodily pain was associated with menstrual pain intensity and premenarcheal MMH [50], suggesting that MMH may represent a risk factor for future pain during development. However, it remains unclear how MMH manifests in adolescence and changes over time. Quantifying MMH longitudinally during adolescence and identifying factors associated with changes in MMH trajectory would help clinicians guide chronic pain management during development [69].

The Early Menstrual Pain Impact on Multisensory Hypersensitivity (EMPATHY) project is the first longitudinal study of MMH development during menarcheal transition. Utilizing a multimodal sensory testing approach, including a non-invasive bladder task, auditory/visual tasks, pain-pressure thresholds (PPTs), conditioned pain modulation (CPM), and cold pain, EMPATHY is uniquely positioned to address questions of MMH development and contributions of menstrual pain to chronic pain trajectories. This investigation had three aims: 1) quantify MMH using a multimodal sensory testing battery; 2) characterize the longitudinal trajectories of MMH in adolescents across the menarcheal transition; 3) identify whether menstrual or non-menstrual pelvic pain is associated with changes in MMH across the menarcheal transition. Given the positive associations between dysmenorrhea, MMH, and chronic pain, we hypothesized that the trajectory of MMH across the menarcheal transition is shaped by exposure to menstrual and pelvic pain.

## Methods

### Study Overview

The study presented herein was part of the larger EMPATHY project (NIH R01HD096332). EMPATHY enrolled premenarcheal adolescents for a three-visit longitudinal study across the menarcheal transition (one premenarcheal baseline visit and two postmenarcheal follow-up visits) to characterize the trajectories of somatic pain, pelvic pain, and dysmenorrhea. Participants were recruited through letters mailed to parents/guardians of age-eligible individuals identified from the Endeavor Health electronic health record. Interested families contacted the study team and were screened for eligibility before enrollment. Comprehensive eligibility criteria are reported elsewhere [50,63] and are listed briefly here. Participants were considered eligible if they were assigned female at birth, between 9–14 years of age, and premenarcheal. Participants were ineligible if they had severe mental health, medical issues, developmental conditions, or language comprehension issues; reported risk factors for prolonged menarche; history of chronic pain; reported use of hormonal suppression; regular use of prescribed analgesics; reported allergies to ultrasound/electrode gel; body mass index > 40kg/m^2^. All participants and their parents provided informed assent and consent to participate, respectively. During each study visit, participants completed questionnaires and experimental tests of MMH. A full study flow diagram of EMPATHY is reported in Osborne et al. [50]. All study procedures adhered to ethical guidelines outlined in the Declaration of Helsinki and received approval from Endeavor Health’s Institutional Review Board.

## Measures

Comprehensive descriptions of all administered assessments are reported elsewhere [50,63]. Brief descriptions of assessments relevant to the presented analyses are included below.

### Questionnaires

Participants completed a Tanner Stage self-Report [45], the Highly Sensitive Child Scale (HSCS) [53], the Children’s Somatic Symptoms Inventory (CSSI) [70], and the short-form Generalized Sensory Sensitivity (GSS) and body map [59]. Each questionnaire was scored according to published guidelines: the HSCS total score comprised the mean of all seven questions and the CSSI total score comprised the summation of all 24 questions, the GSS comprised the summation of six possible symptoms, and the body map comprised the summation of seven possible body sites. Menstrual pain was assessed using a numerical rating scale (NRS) from 0 (no pain) – 10 (worst pain imaginable) in response to the question: “Over the last 3 months, what was the average cramping pain you experienced with your period (around or below your belly button or lower back)?” Pelvic pain was the average NRS rating in response to the following three questions: 1) “Over the last 2 weeks, how much pelvic pain (NOT including any period pain) have you experienced?”; 2) “Please indicate over the last 2 weeks how much pain you have experienced when having a bowel movement.”; 3) “Please indicate over the last 2 weeks how much pain you have experienced when emptying your bladder.”

### Tests of MMH

The non-invasive bladder filling task [64] measured participants’ bladder pain and urgency ratings using a 0–100 visual analog scale (VAS) at four timepoints. After an initial bladder void, each participant rated their baseline bladder pain and urgency and then drank 20 fl. oz. of water. Participants reported when reaching first sensation (FS; “you are aware there is some urine in your bladder”), first urge to urinate (FU; “you could go to the bathroom to pee, but you do not have to rush off to do this”), and maximum tolerance (MT; “you would leave in the middle of your favorite movie to go to the bathroom even if you were missing the best part”). Pain and urgency ratings were obtained for each sensation, and if maximum tolerance was not reached within two hours, the two-hour ratings were used. Participants voided once reaching maximum tolerance.

Pain pressure thresholds (PPT) were assessed at the right shoulder and knee using a digital Algometer (Wagner Instruments, Greenwich, CT) with 1 cm^2^ tip. At each site, pressure was increased via software generated ramp at 4 N/s until the participant first reported the sensation as painful via a joystick. Two trials were obtained per site, and the average of these two trials served as the PPT. If the trial was invalid (e.g., equipment malfunction), the single valid trial was used. After each PPT trial, participants rated their after-pain using an NRS. A third after-pain rating for the shoulder and knee was collected after conditioned pain modulation (CPM).

To obtain a measure of CPM, participants first completed a 20 second cold pressor task by submerging their dominant hand in water chilled to 7±2 C° using ice and a circulating water pump to maintain consistent temperature [64]. During the cold pressor, cold pain NRS ratings of the submerged hand were collected at 10 and 20 seconds. Immediately after the cold pressor, a knee PPT measurement was obtained. Subtracting the averaged knee PPT measure, which was completed earlier in the study visit, from this post-cold pressor knee PPT measure served as our CPM measure.

Sensitivity to visual stimuli was assessed by presenting participants with a blue/yellow pattern-reversal (25Hz) checkerboard stimulus on a computer screen in a dark room for 20 seconds across five blocks of different brightness intensities (1, 30, 60, 90, or 120 lux). Following each block, participants rated their perceived unpleasantness using the Gracely Box Scale that increments from 0–20 with verbal descriptors [21]. The auditory task was a similar design, except participants listened to a tone presented in pneumatic earphones at randomly presented loudness intensities (15, 30, 45, or 60 dB). After each 20 second block, participants rated their perceived unpleasantness using the Gracely Box Scale. Scalp electroencephalography (EEG) was collected during visual/auditory stimulation but was not used in the present investigation. EEG results are reported elsewhere [50]. For both visual and auditory tasks, the average unpleasantness rating and the slope between stimulus intensity and unpleasantness (i.e., stimulus response function) were calculated.

## Procedure

Upon enrollment, eligible participants completed an in-person premenarche baseline visit and two postmenarche visits (separated by one year) at Evanston Hospital (Evanston, IL). Study visits comprised questionnaires during the bladder task and experimental tests of MMH afterwards.

## Statistical Analysis

### Quantification of MMH

A total of 24 measurements from the experimental pain and sensory sensitivity assessments were entered into a principal component analysis (PCA): stimulus-response slopes and mean unpleasantness ratings from visual and auditory tasks (4); pain and urgency ratings from the bladder task at baseline, FS (green zone), FU (yellow zone), and MT (red zone) (8); shoulder and knee PPTs (2); after-pain ratings from the two knee and shoulder PPT trials and following the cold pressor (6); knee CPM (1); cold pain ratings at 10s and 20s, and hand after-pain following the cold pressor task (3).

During the bladder task, some participants failed to provide pain and urgency ratings across the full spectrum of zones, either by skipping first sensation or never reaching first urge by the specified time limit. Therefore, missing pain and urgency ratings were estimated using multiple imputation by chained equations [10]. Across all longitudinal timepoints, a total of *n*=299 participants had at least two observations of pain or urgency ratings during the bladder task within each session and therefore were eligible for imputation. A two-level predictive mean matching (PMM) approach was implemented separately for pain and urgency variables, using the shared experimental stage as a predictor and treating subject-visit combinations as cluster variables. Twenty imputed datasets were generated with a single iteration using a fixed random seed for reproducibility. The imputation procedure preserved the longitudinal structure of the data by treating each participant-visit combination as independent units. The percentage of data points imputed in the total sample (*n*=299) was 10% (both pain and urgency) at baseline, 13% (both pain and urgency) at the first postmenarcheal visit, and 12% (both) at the second postmenarcheal visit. The percentage of bladder pain and urgency ratings imputed for subsequent PCA and longitudinal modeling datasets were very similar (within 2%).

Prior to the PCA, data were mean centered and normalized to unit variance (i.e., z-scored). To standardize all measurements such that an increase in value denoted an increase in pain/sensory sensitivity/impairment, the knee PPT and shoulder PPT and CPM measures were multiplied by -1. Only participants with complete data across all 24 measures at their baseline visit were entered into the PCA. The PCA was computed using singular value decomposition [2]. When interpreting the PCs, we employed permutation testing of the PCs, bootstrapping to identify which variables significantly contributed to the variance of each PC via bootstrap ratios, and geometric plotting of loadings [3,6]. Both permutation testing and bootstrapping procedures used 2,000 iterations with a fixed random seed for reproducibility. Loadings of the four questionnaires were estimated via supplementary projections onto the already-established PC space [6]. Importantly, these questionnaires were not used in the initial PCA model training; rather, they were projected into the existing PC space to examine their relationships with the experimentally derived components. We computed correlations between PC factor scores and questionnaire outcomes, and adjusted for multiple comparisons by controlling the false discovery rate (*p*_fdr_<0.05) [7]. As with any empirically derived component solution, the specific loadings and factor weights reported here are sample-specific; we discuss the generalizability of the pattern versus the weights in the Discussion.

Longitudinal factor scores for each participant’s second and third visit were also estimated via supplementary projections using the same baseline-trained PCA model. Using supplementary projections to estimate longitudinal changes in PCs provided the advantage of imposing the same multivariate structure across all participants, which was constructed with maximal data collected at baseline, and represents a multivariate space prior to menarche where MMH is hypothesized to be stable.

### Longitudinal Modeling of MMH

Once we identified a PC of MMH using PCA, we estimated its change over time using linear mixed modeling. Predictors included menstrual pain and pelvic pain reported at the participants’ second visit (i.e., first postmenarche visit), time since baseline in years, and their interactions. Tanner stages (breast and pubic hair) recorded at each visit were entered as covariates to adjust for pubertal development. To increase interpretability of the intercept term, Tanner stages were averaged within each participant and grand mean centered across the sample. The difference in Tanner stage at each visit relative to each participant’s mean Tanner stage served as the within-subject change in pubertal development. Therefore, Tanner staging was modeled as mean-centered between-subject and within-subject changes in accordance with best practices [16,28]. Menstrual and pelvic pain were likewise grand mean centered. Time was centered at 1.57 years, the mean interval from the baseline visit to the first postmenarcheal visit, at which menstrual and pelvic pain were reported. The menstrual and pelvic pain coefficients therefore describe their association with MMH at the occasion when pain was measured. A mixed model with random effects of participant-wise intercepts and slopes of time was compared to a simpler random intercept model using analysis of variance (ANOVA).

### Attrition Sensitivity Analyses

Because only 119 of the 214 baseline-PCA participants (55.6%) contributed to the longitudinal models, we conducted a set of attrition sensitivity analyses, reported in full in the Supplemental Material (note that *n*=140 total participants were included in longitudinal modeling; therefore, the remaining 21 mixed-model participants entered via follow-up visits only). We compared retained and non-retained participants across all 24 PCA input measures, four questionnaires, and demographics using Welch’s *t*-tests and quantified effect sizes using Hedges’ *g*; tested invariance of the factor structure across retention groups using Procrustes-aligned Tucker congruence [41,57] and *R_V_* coefficients [1] benchmarked against a 2,000-split permutation null; and evaluated whether attrition distorted the estimated developmental trajectory using a pattern-mixture extension of the mixed model—allowing the developmental trajectory to vary by attrition pattern (completers vs. dropouts)—and inverse-probability-of-retention weighting [29].

### Precision and Measurement Reliability

Sample size was determined by the parent EMPATHY study (R01HD096332), whose *a priori* power analysis was designed for a structural equation model with a planned enrollment of 375 adolescents. Enrollment was curtailed by the COVID-19 pandemic, yielding 300 enrolled participants, of whom 214 had complete baseline data, and 140 contributed to the longitudinal models. Because the present analysis used linear mixed-effects models rather than the originally planned SEM, we conducted a simulation-based sensitivity analysis to characterize the effects detectable in the realized sample (see Supplementary Material).

Because menstrual and pelvic pain differed substantially in dispersion, their per-unit coefficients were not directly comparable. We therefore tested the difference between them on a common scale by rescaling each coefficient by the subject-level standard deviation of its predictor and evaluated the linear contrast (pelvic − menstrual) against the fitted model’s fixed-effect covariance matrix, with Satterthwaite degrees of freedom.

Menstrual pain was assessed as a recall-based rating of average cramping pain over the preceding three months, a format susceptible to measurement error that would attenuate its association with MMH. We therefore estimated its test–retest reliability across the two postmenarcheal visits using a two-way random-effects, absolute-agreement, single-measure intraclass correlation coefficient (ICC2) among participants in the estimation sample who provided ratings at both visits (*n*=81), and applied a classical correction for attenuation to the menstrual pain coefficient.

All statistical analyses were conducted in R (v.4.4.1) [54] using the *mice* package for multiple imputation [10], the *ExPosition* family of R packages for PCA with inferential statistics [6], *lme4* [5] and *lmerTest* [36] for linear mixed modeling, Satterthwaite degrees of freedom, and the linear contrast between pain coefficients. Standardized coefficients were computed with the *parameters* package [42], intraclass correlations with *performance* [43] (unconditional model) and *psych* [56] (test–retest), and the simulation-based sensitivity analysis with *simr* [22]; minimum detectable effects were obtained as the 80% effective dose of a probit model fitted to the simulated rejection rates using *MASS* [66]. Data processing and visualization used the *tidyverse* packages [74].

## Results

From the 300 participants who enrolled in EMPATHY, 214 had complete data across all MMH testing measures at baseline (see Figure 1). Participants eligible for longitudinal modeling were required to have complete MMH testing variables for at least one visit and have completed the menstrual and pelvic pain questions at their postmenarche visit 1; therefore, a total of 140 participants were eligible for longitudinal modeling. Baseline demographics and measures for the baseline PCA and longitudinal cohorts are shown in Table 1. At baseline, the cohort was characterized as slightly older than 11 years of age, majority white/caucasian, not Hispanic or Latino, and from affluent households; characteristic of the communities surrounding Evanston Hospital where this study was located.

**Figure 1.**
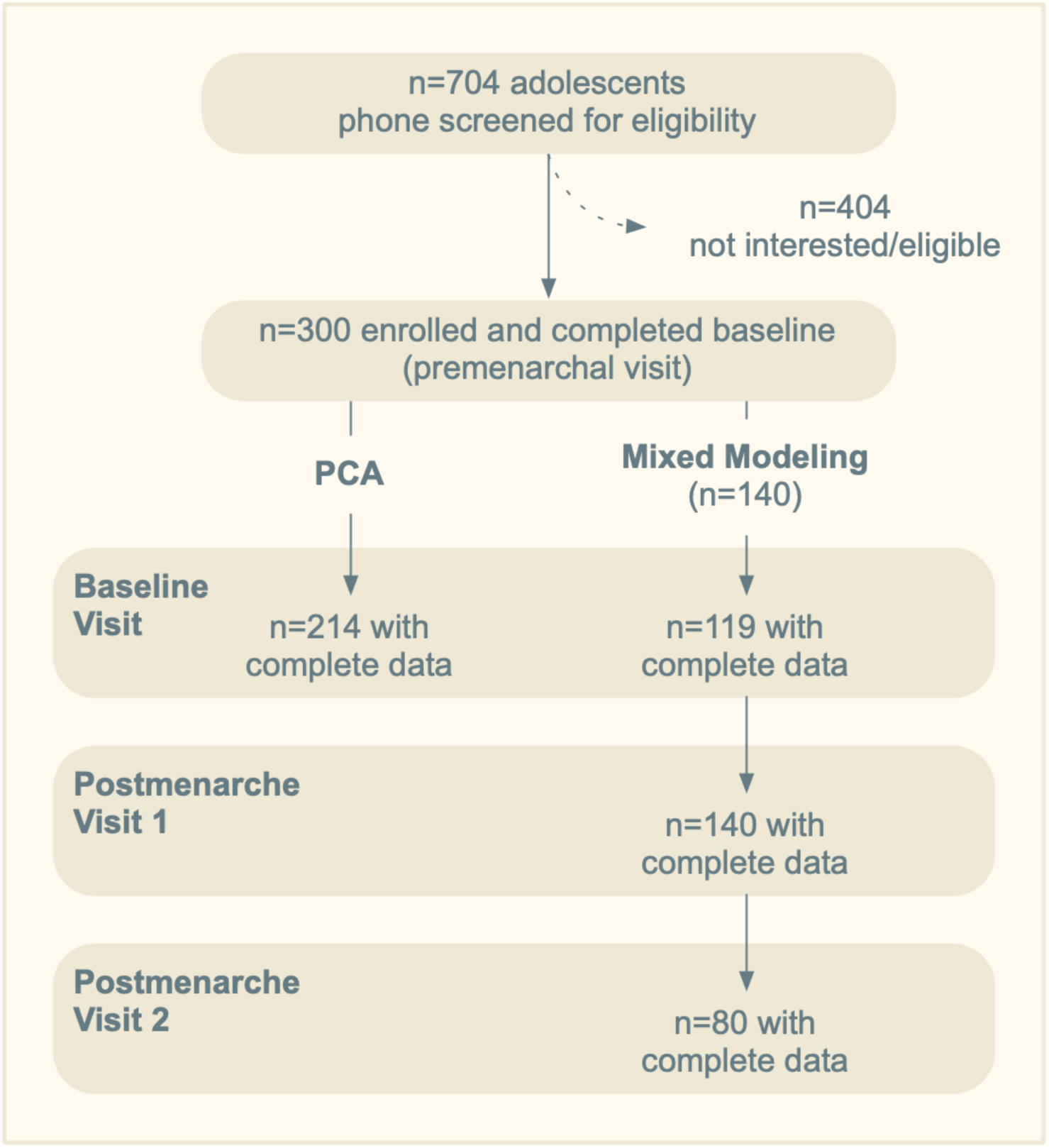
EMPATHY participant flow diagram. After screening and enrollment, participants completed a baseline premenarcheal visit, and two postmenarcheal visits. Postmenarcheal visit 2 occurred ∼1 year following the first postmenarcheal visit. The principal component analysis (PCA) quantifying multimodal hypersensitivity (MMH) was performed with all eligible participants with complete baseline data. Linear mixed-effects modeling similarly used all available longitudinal data; therefore, participants lost to follow-up were included, minimizing bias.

**Table 1.** Participant Baseline Demographics by Analysis.

| Category | Measure | Analysis Cohort |  |
| --- | --- | --- | --- |
|  |  | PCA | Mixed-Modeling |
| Sample Size |  | 214 | 140 |
| Age (years) | Age | 11.29 (0.98) | 11.31 (0.87) |
| Race | American Indian or Alaska Native | 3 (1.4%) | 3 (2.14%) |
|  | Asian | 28 (13.08%) | 22 (15.71%) |
|  | Black or African American | 15 (7.01%) | 9 (6.43%) |
|  | Native Hawaiian or Other Pacific Islander | 1 (0.47%) | 1 (0.71%) |
|  | White or Caucasian | 190 (88.79%) | 121 (86.43%) |
| Ethnicity | Hispanic or Latino | 12 (5.61%) | 5 (3.57%) |
|  | Non-Hispanic or Latino | 202 (94.39%) | 135 (96.43%) |
| Income | <\$25k | 3 (1.4%) | 1 (0.71%) |
| | \$25k - <\$50k | 7 (3.27%) | 5 (3.57%) |
| | \$50k - <\$75k | 10 (4.67%) | 6 (4.29%) |
| | \$75k - <\$100k | 20 (9.35%) | 16 (11.43%) |
| | \$100k - <\$150k | 37 (17.29%) | 29 (20.71%) |
| | ≥\$150k | 126 (58.88%) | 75 (53.57%) |
|  | Prefer not to answer | 11 (5.14%) | 8 (5.71%) |
| Tanner Stage (Breast) | 1 | 23 (10.75%) | 11 (7.86%) |
|  | 2 | 87 (40.65%) | 49 (35%) |
|  | 3 | 84 (39.25%) | 63 (45%) |
|  | 4 | 17 (7.94%) | 14 (10%) |
|  | 5 | 2 (0.93%) | 3 (2.14%) |
|  | Missing | 1 (0.47%) | 0 (0%) |
| Tanner Stage (hair) | 1 | 29 (13.55%) | 12 (8.57%) |
|  | 2 | 95 (44.39%) | 61 (43.57%) |
|  | 3 | 65 (30.37%) | 46 (32.86%) |
|  | 4 | 22 (10.28%) | 19 (13.57%) |
|  | 5 | 2 (0.93%) | 2 (1.43%) |
|  | Missing | 1 (0.47%) | 0 (0%) |

*Note.* Categorical measures are presented as *n* (%); age is presented *M* (*SD*). PCA=principal component analysis.

## Quantification of MMH

We performed a PCA on participants with complete data from their baseline visit (*n*=214). The scree plot with permutation testing results indicated that the first six (out of 24) PCs were significant (see Figure S1). We further interrogated the first six PCs by examining geometrically plotted factor score loadings and comparing bootstrap ratios across the measures. We determined that the first three PCs, which comprised 34.2% of the variance, were distinct and interpretable components of pain processing and sensory sensitivity from our experimental testing battery. All experimental measures loaded onto PC1 (σ^2^=16.49%, *p*=0.0005) with significant bootstrap ratios (see Figure 2), suggesting positive associations across all pain and sensory sensitivity measures (see Figure 3A). Plotting standardized MMH measures similarly demonstrated separation between low versus high MMH participants derived via quartiles (see Figure S2). Therefore, we interpreted PC1 as representing MMH.

**Figure 2.**
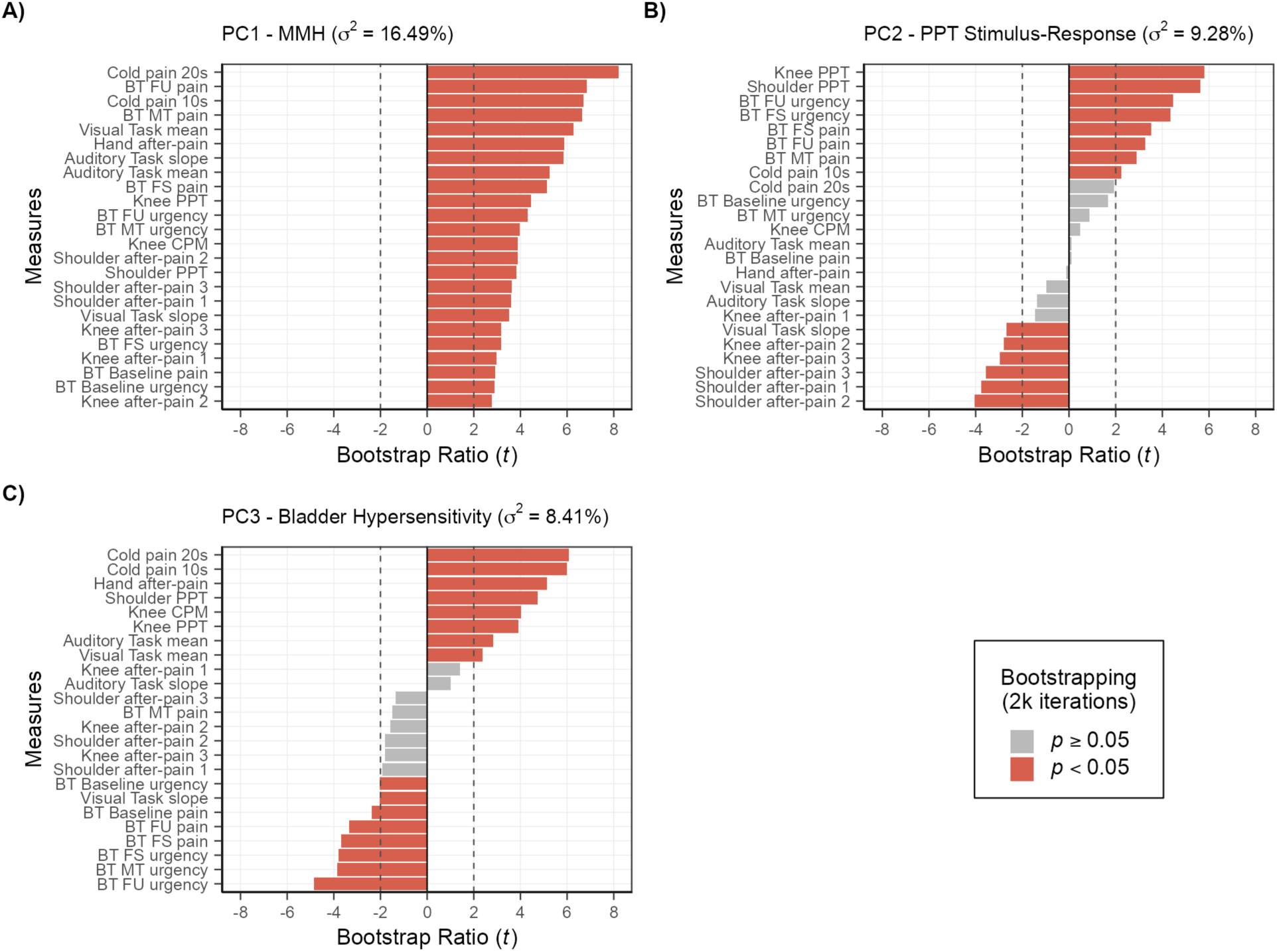
Bootstrap ratios of loadings. A) All measures loaded positively on principal component 1 (PC1) with significant bootstrap ratios, indicating that all measures contributed to PC1’s variance. Given the positive associations with the measures to each other, we interpret PC1 as multimodal hypersensitivity (MMH). B) Pressure pain testing (PPT) measures were diametrically opposed to their corresponding after-pain measurements on PC2, indicating a relationship between applied algometer force and reported after-pain (i.e., less force applied; less pain reported). We interpret this component as the PPT stimulus-response function. C) PC3 was characterized by a clustering of measures from the non-invasive bladder pain task, suggesting PC3 as representing bladder hypersensitivity.

**Figure 3.**
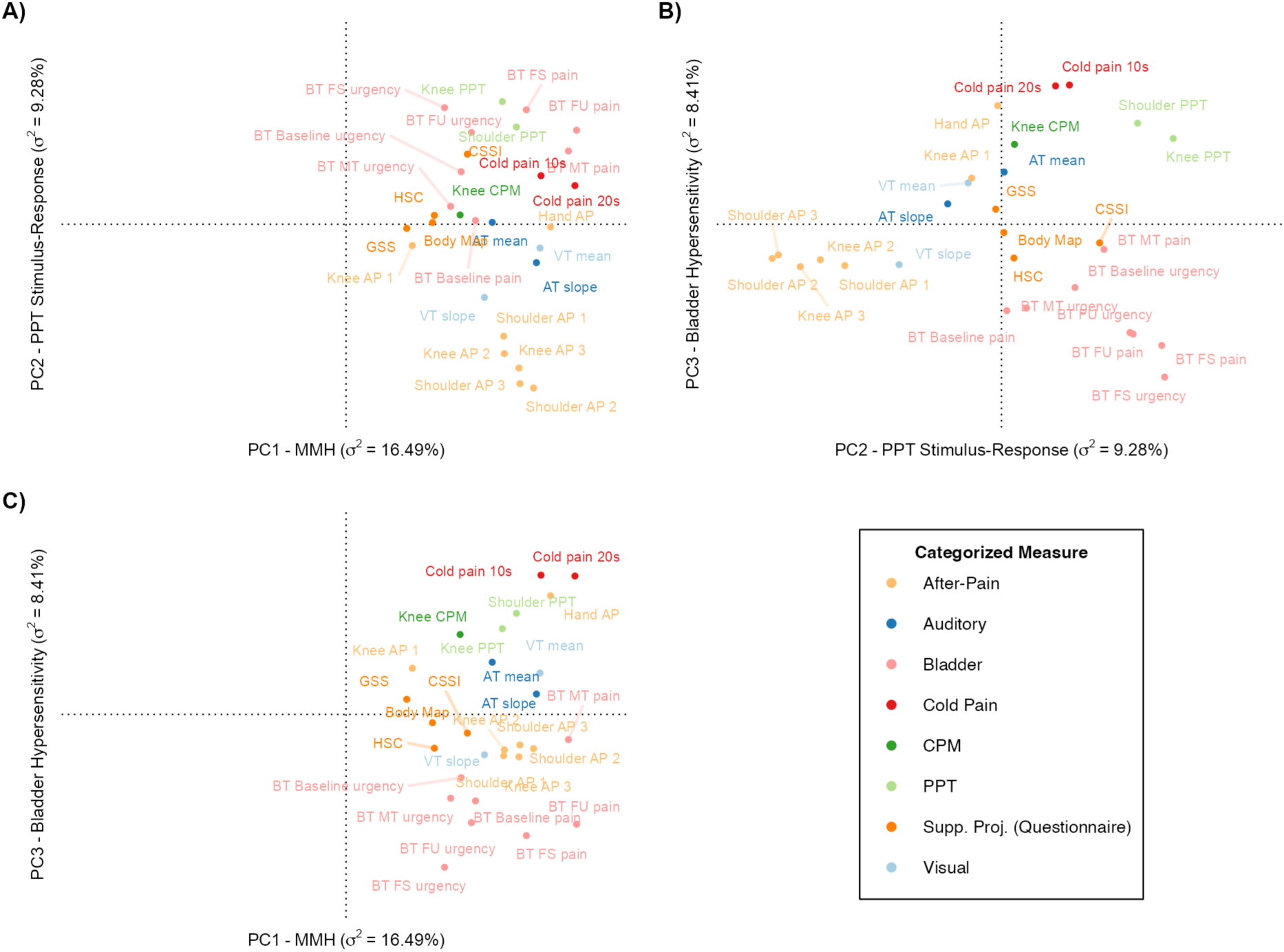
Geometric scatter plots of factor loadings from PC1–3. A) All experimentally derived measures of sensory sensitivity load to the right of the origin on PC1 (x-axis), indicating a positive relationship between sensitivity across all measures, i.e., multimodal hypersensitivity (MMH). Symptom questionnaires of sensory sensitivity were projected onto the multivariate space and also load onto PC1. B) When plotting PC2 (x-axis) against PC3 (y-axis) a clustering of bladder pain task measures is evident in the fourth quadrant, indicating bladder hypersensitivity represented on PC3. PC2 opposes PPTs with their corresponding after-pain measurements, indicating a PPT stimulus-response function. C) Plotting PC1 (x-axis) against PC3 (y-axis) provides another perspective, contrasting bladder pain testing measures with peripheral measures—cold pain, CPM, and PPT measures—on PC3. BT=bladder task; FS=first sensation; FU=first urge; MT=maximum tolerance; PPT=pain pressure threshold; AP=after-pain; AT=auditory task; VT=visual task; GSS=generalized sensory sensitivity; HSC=highly sensitive child; CSSI=children’s somatic symptoms inventory; CPM=conditioned pain modulation; PC=principal component.

A negative relationship between PPT measures and their respective after-pain ratings was observed across PC2 (σ^2^=9.28%, *p*=0.0005) and orthogonal to PC1 MMH (see Figure 3B). Deploying more force/pressure during PPT testing was associated with greater after-pain ratings. We interpreted PC2 as the mechanical stimulus-response function of PPT testing (PPT-SR).

Across PC3 (σ^2^=8.41%, *p*=0.0005), we observed the clustering of the bladder test and measures collected during CPM testing—knee CPM and ratings of pain during and after cold pressor—across the origin (see Figure 3C). This negative association between bladder test pain/urgency ratings and CPM testing measures may reflect participants’ differential sensitivity to peripheral versus visceral stimulation that is orthogonal to MMH and the PPT S-R.

## Relationship between MMH and Symptom Questionnaires

We performed a sensitivity analysis of the first three PCs by examining how four self-report questionnaires examining differing aspects of sensory sensitivity would load onto the previously established PC space via supplementary projections. This supplementary projection approach allowed us to examine relationships between questionnaires and PCs without altering the original factor structure, ensuring that the experimental measures remain the primary determinants of the PC space and MMH.

All four sensory sensitivity questionnaires loaded on PC1 (see Figure 3), showing positive associations with all experimental measures. Additionally, the participants’ factor scores on PC1 positively correlated with the scores from each questionnaire: Body Map, *r*(212)=0.211 [0.079, 0.336], *p*_fdr_=.006; CSSI, *r*(212)=0.296 [0.169, 0.414], *p*_fdr_=1.26x10^-4^; HSC, *r*(212)=0.216 [0.084, 0.340], *p*_fdr_=.006. However, the GSS did not survive correction for multiple comparisons: *r*(212)=.15 [0.014, 0.277], *p*_fdr_=.073. Together, these findings further support the interpretation of PC1 as MMH.

The pattern of questionnaire supplementary projection loadings was less pronounced for the other components (see Figure 3) and questionnaire scores did not correlate with participant factor scores except for the CSSI. The CSSI loaded onto PC2, and demonstrated a positive correlation with participant factor scores on PC2, *r*(212)=.24 [0.109, 0.362], *p*_fdr_=.002, indicating that increased sensory sensitivity as measured by the CSSI was associated with more sensitive PPTs (orthogonal to MMH).

## Longitudinal MMH Change and Associations with Menstrual and Pelvic Pain

A total of *n*=140 participants had complete data for longitudinal modeling: *n*=9 (6%) participants contributed data from only one visit, *n*=63 (45%) two visits, and *n*=68 (49%) all three visits (see Figure 1). The average number of years since baseline was 1.57 years (*SD*=0.79) for postmenarcheal visit 1 and 2.66 years (*SD*=0.70) for postmenarcheal visit 2.

We observed that the linear mixed model with random intercepts and slopes of time (AIC=1288.5) resulted in an improved model fit, χ^2^(2)=8.68, *p*=0.013, compared to the random intercept-only model (AIC=1293.2), suggesting individual variability in MMH trajectories across the menarcheal transition. Therefore, we present the more complex random intercept and slope model herein (see Table 2). Variance in MMH was predominately between-person: the intraclass correlation coefficient (ICC) from an unconditional random-intercept model was 0.585, indicating that 58.5% of the variance in MMH was between participants and 41.5% within participants across visits. MMH is therefore more trait-like than state-like across this interval.

**Table 2.** Linear Mixed-Modeling Regression Results.

| Term | *b* (95% CI) | *SE* | *df* | *t* | *p* | $\beta$ (95% CI) |
| --- | --- | --- | --- | --- | --- | --- |
| Intercept | -0.225 (-0.485, 0.035) | 0.133 | 130.929 | -1.698 | 0.092 | 0.023 (-0.119, 0.164) |
| Tanner Stage - Breast (Between-Subject Effect) | 0.028 (-0.373, 0.429) | 0.205 | 115.243 | 0.137 | 0.891 | 0.010 (-0.129, 0.149) |
| Tanner Stage - Breast (Within-Subject Change) | -0.002 (-0.264, 0.261) | 0.134 | 206.770 | -0.014 | 0.989 | -0.001 (-0.104, 0.103) |
| Tanner Stage - Hair (Between-Subject Effect) | -0.240 (-0.625, 0.144) | 0.196 | 131.990 | -1.227 | 0.222 | -0.087 (-0.226, 0.052) |
| Tanner Stage - Hair (Within-Subject Change) | -0.042 (-0.290, 0.206) | 0.127 | 220.227 | -0.331 | 0.741 | -0.019 (-0.134, 0.095) |
| **Time** | **-0.229 (-0.434, -0.025)** | **0.104** | **183.896** | **-2.199** | **0.029** | **-0.146 (-0.278, -0.015)** |
| Menstrual Pain | 0.070 (-0.038, 0.178) | 0.055 | 125.553 | 1.274 | 0.205 | 0.074 (-0.071, 0.220) |
| **Pelvic Pain** | **0.644 (0.397, 0.892)** | **0.126** | **123.308** | **5.101** | **1.24e-06** | **0.415 (0.256, 0.573)** |
| Time * Menstrual Pain | 0.041 (-0.013, 0.094) | 0.027 | 88.236 | 1.494 | 0.139 | 0.063 (-0.021, 0.146) |
| Time * Pelvic Pain | -0.041 (-0.159, 0.078) | 0.061 | 88.902 | -0.672 | 0.503 | -0.031 (-0.120, 0.058) |
| Menstrual * Pelvic Pain | -0.022 (-0.121, 0.077) | 0.050 | 118.219 | -0.444 | 0.658 | -0.031 (-0.187, 0.126) |
| Time * Menstrual * Pelvic Pain | -0.008 (-0.052, 0.036) | 0.022 | 60.842 | -0.355 | 0.724 | -0.015 (-0.095, 0.066) |

*Note.* Between-subject effects for Tanner stage were grand mean centered across participants; within-subject change reflects deviation from each participant's mean Tanner stage. Menstrual and pelvic pain were grand mean centered, and time was centered at the mean interval of the visit at which menstrual and pelvic pain were assessed, so each unstandardized coefficient is the effect at sample-average levels of the other predictors and at that occasion. Standardized coefficients were estimated by refitting the model after z-scoring all continuous predictors and the outcome. *b*=unstandardized coefficient; CI=confidence interval.

We observed a decrease in MMH over time (*b*=-0.229, β=-0.15, *p=*0.029) and a positive association between MMH and pelvic pain (*b*=0.644, β=0.42, *p*=1.24x10^-6^; see Figure 4). There was no association observed between MMH and menstrual pain (*p*=0.205), and time did not interact with menstrual pain (*p*=0.139) or pelvic pain (*p*=0.503).

**Figure 4.**
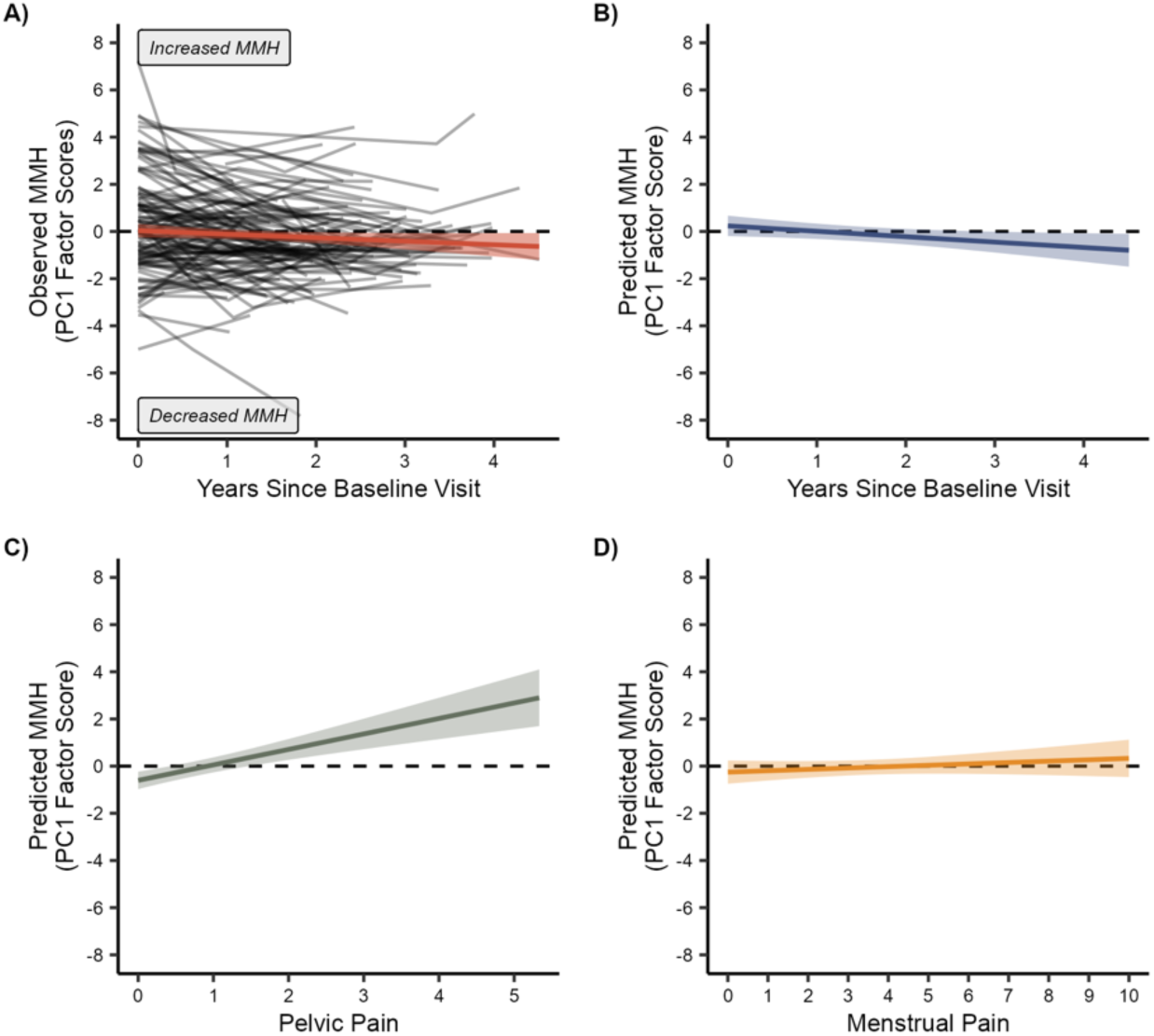
Longitudinal MMH is associated with pelvic pain and not menstrual pain. A) Observed longitudinal trajectories of MMH are plotted for each participant in black and study-wise average slope in red. B) Predicted MMH change over time is plotted using linear mixed-modeling coefficients at sample averages for pubertal development, menstrual pain, and pelvic pain. C) Predicted MMH as a function of pelvic pain at sample averages for pubertal development, menstrual pain, and follow-up time. D) Predicted MMH as a function of menstrual pain at sample averages for pubertal development, pelvic pain, and follow-up time. Panels B–D use the sample’s observed min. to max. value of the x-axis. Shading denotes 95% CI.

To visualize the relationship between pelvic pain and MMH, we split participants into three groups. The “no pain” group comprised participants reporting zero pelvic pain (*n*=65). The remaining participants were split across median pelvic pain, resulting in “some pain” (0.33–1.33; *n*=41) and “more pain” (1.67–5.33, *n*=34) groups. Similarly, we predicted MMH as a function of the median score within these groups over time at average menstrual and pubertal development (see Figure 5B). Participants with the most pelvic pain (“more pain” group) demonstrated elevated observed and predicted MMH. In sum, after adjusting for time and pubertal development, MMH was associated with pelvic pain, and not menstrual pain.

**Figure 5.**
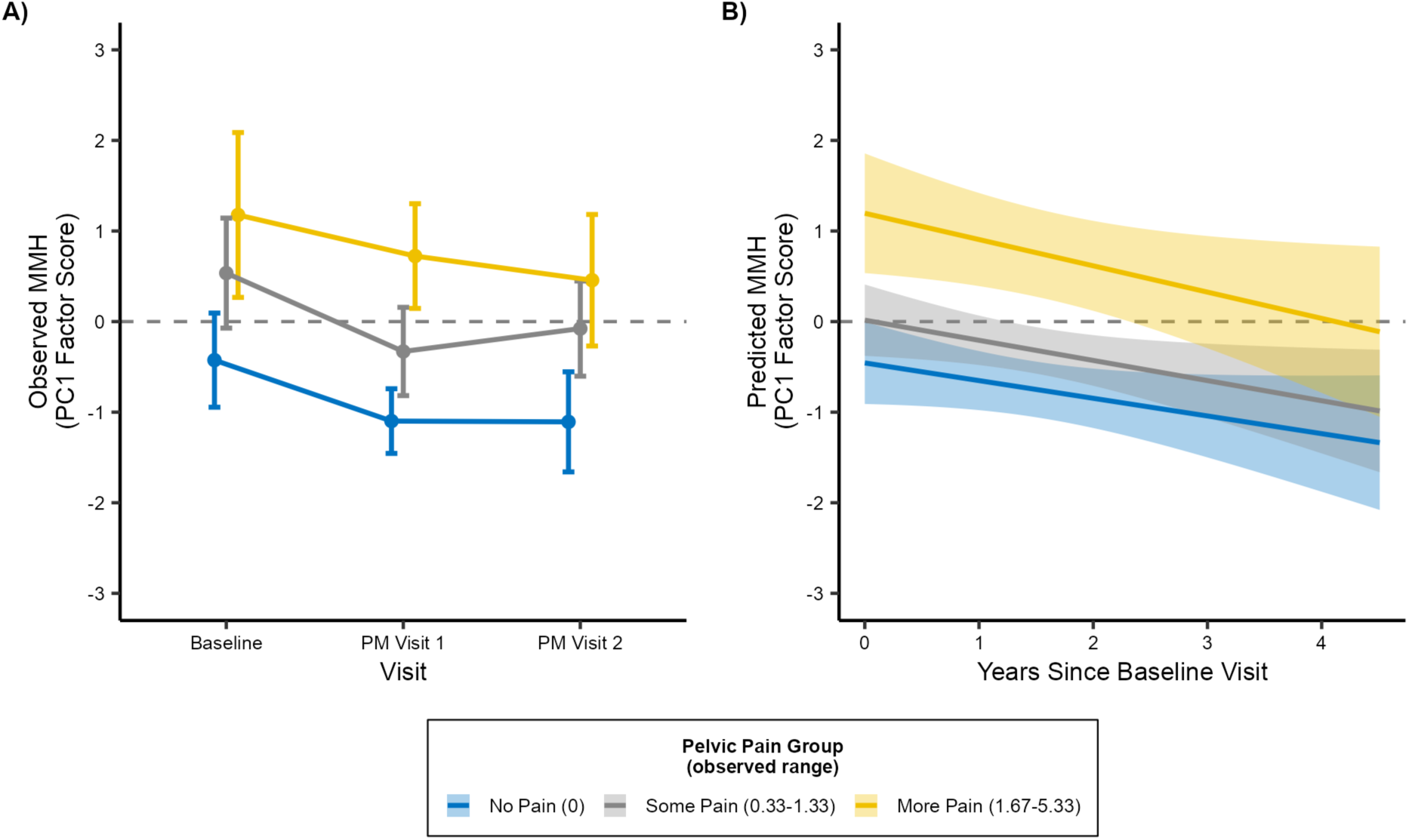
Pelvic pain groups and longitudinal MMH. A) Participants split into groups of no, some, and more pelvic pain demonstrated a consistent pattern of elevated observed MMH with increased pelvic pain. B) Predicting MMH in the typical participant within each pelvic pain group over time, while adjusting for menstrual pain and pubertal development, also demonstrated a pattern of elevated MMH over time. The figure legend denotes the observed range of mean pelvic pain NRS in parentheses. Menstrual and pelvic pain were derived from participants’ first postmenarche visit. Error bars and shading denote 95% CIs.

## Precision and Measurement Reliability and Robustness to Attrition

The null menstrual pain association was not attributable to insufficient sensitivity or unreliable measurement (see Supplemental Materials for full analyses). The upper confidence limit bounds it at 28% of the pelvic pain estimate, detectable effect sizes were smaller per NRS point for menstrual than for pelvic pain (see Table S1), and a direct per-SD contrast confirmed the larger pelvic association (*p*=0.016). Menstrual pain also showed moderate test–retest reliability (*ICC2*=0.53). The results are unlikely biased due to attrition: retained and non-retained participants differed at baseline only in pubertal stage and modestly higher baseline MMH among those retained, the MMH factor structure was invariant to retention (see Table S2), and neither pattern-mixture nor inverse-probability-weighted models materially altered the pelvic pain association (see Table S3).

## Discussion

In a longitudinal study examining MMH across the menarcheal transition in female adolescents, we observed three novel findings. First, and to our knowledge for the first time in adolescents, we showed that MMH can be derived as a single multivariate dimension integrating experimental measures of pain and sensory sensitivity. Second, participants’ MMH changed over time, decreasing on average with each subsequent visit. Third, greater pelvic pain, but not menstrual pain, was associated with greater MMH after adjusting for time and pubertal development.

In a PCA of all experimental and QST measures encompassing CPM and multiple sensory modalities (mechanosensation, thermosensation, vision, audition, visceral pain and urgency), we observed three main PCs: MMH (PC1), PPT stimulus-response function (PC2), and bladder hypersensitivity (PC3). A similar MMH testing procedure in a cohort of young adult women identified nearly identical components [33], suggesting that these components are robust across age and pubertal development in females within this experimental testing protocol. We observed robust converging evidence suggesting that PC1 reflects participant MMH: 1) significant bootstrap ratios were observed across all measures on PC1 and 2) all questionnaires of sensory sensitivity (i.e., patient-reported outcomes) loaded onto the MMH component as supplementary projections (i.e., untrained data) and correlated with participant MMH factor scores. Taken together, this study replicated our findings in an adolescent cohort, provided confidence that PC1 isolates MMH, and contributed a multimodal QST study in adolescents, a population dominated by single-modality measures [65]. Beyond our prior adults study [33], another factor-analytic work in experimental pain in adults was consistent with a component reflecting variance shared across sensory measures: a PCA of thermal, mechanical, and electrical pain thresholds recovered a dominant first component loading on all stimuli [49]. However, another large QST dataset reported several separate modality-specific components [23].

Critically, both were restricted to nociceptive stimuli and did not include non-pain modalities such as visual unpleasantness or visceral urgency, leaving open whether such a shared-variance component generalizes across pain and non-pain sensory domains.

The pre- to postmenarcheal longitudinal modeling of MMH demonstrated that MMH was highest at the participants’ baseline visit and, on average, decreased over time; notably, the random effect of time demonstrated reliable differences in MMH trajectories across participants (see Figure 4A). A decrease in sensitivity with age in children and adolescents was previously observed in mechanical and thermal pain thresholds [9] and cold pain [27], but not in non-nociceptive measures. One potential explanation is the maturation of central pain inhibitory mechanisms, as older children exhibit greater CPM than younger children [61]. To our knowledge, this is the first study to quantify longitudinal changes in MMH in a multivariate manner.

Contrary to our motivating hypothesis, we did not observe a relationship between menstrual pain and MMH. Rather, pelvic pain was positively associated with MMH independent of participants’ age, pubertal development, and menstrual pain. Participants reporting greater pelvic pain consistently had greater MMH across multiple years. This aligns with studies in adults with urologic chronic pelvic pain syndromes that have shown increased somatosensory sensitivity in areas outside the pelvis [12,25]. Pelvic pain, and not menstrual pain, was associated with visual hypersensitivity [34], implicating supraspinal sensory sensitization.

However, in the presented analysis pelvic pain was assessed at participants’ first postmenarcheal visit, whereas MMH was measured from the premenarcheal baseline onward. Because the time × pelvic pain interaction was not significant (*p*=0.503), the association between pelvic pain and MMH is better characterized as a stable between-person level difference than as a pain-driven change in trajectory. In other words, we observed a trait-like elevation in pain system sensitivity that co-varies with pelvic pain reporting across time. Whether such elevations reflect repeated nociceptive exposure altering central processing or instead index pre-existing differences in central nervous system sensitivity remains a long-standing question in dysmenorrhea research [8]; our non-significant time interaction is most consistent with the latter. This finding converges with Osborne et al. [50] who demonstrated that premenarcheal MMH prospectively predicts postmenarcheal widespread pain. That a single pre-menarche phenotype associates with two distinct post-menarche pain outcomes within the same cohort strengthens the case for MMH as a genuine antecedent risk marker of nociplastic pain. This interpretation aligns with prospective cohort data in adults, in which heightened somatic and sensory complaint precedes and predicts incident temporomandibular disorder [17] and chronic widespread pain [46], and in which non-pelvic somatic symptom burden tracks with urologic chronic pelvic pain [35]. This stable co-variation of MMH is consistent with MMH functioning as an early marker of nociplastic pain [18]. Because neuroplasticity is heightened during early development, which allows adverse sensory experiences to shape later pain sensitivity [60,67,72], characterizing how such a phenotype is established and maintained across the perimenarcheal transition has direct implications for identifying intervention windows before nociplastic pain becomes clinically entrenched.

Given our adolescent cohort, less exposure to painful menses is one potential explanation for the lack of relationship between menstrual pain and MMH. Primary dysmenorrhea, painful menses without anatomical pathology, is a known risk factor for chronic pain conditions [30,40,55]. One proposed mechanism underlying this risk is central sensitization driven by repeated cyclical exposure to menstrual pain [30,32], although whether dysmenorrhea itself meets criteria for a central sensitization syndrome remains debated [62]. One possibility is that the duration of menstrual pain exposure in our cohort (≤ ∼3 years postmenarche) was insufficient to drive detectable changes in MMH, whereas the young adult cohort in Kmiecik et al. [33] had a longer exposure window. Consistent with a cumulative-exposure account, primary dysmenorrhea tends to remain relatively stable across early adulthood [73]. Relative to women with less exposure, women with long-term dysmenorrhea reported greater areas of pain sensitivity during menses and following pain-pressure stimulation [19]. Monitoring exposure to menstrual pain in conjunction with non-menstrual pelvic pain through pubertal development may shed more light on how these pain mechanisms may interact to modulate MMH and increase risk of chronic pain.

The co-occurrence of elevated MMH and pelvic pain, present even at premenarcheal baseline, emphasizes the importance of early screening to identify adolescents at risk for both conditions across pubertal development [51,69]. Although whether interventions targeting pelvic pain in adolescence can reduce MMH remains untested, indirect evidence from pediatric and adult studies suggests this is a promising avenue. In children with unexplained abdominal pain, cognitive-behavioral therapy improved pain and gastrointestinal symptoms relative to a control treatment [39,47]. In adults, treatment of non-menstrual visceral pain conditions, including dietary management of IBS, surgical treatment of symptomatic gallbladder calculosis, and management of diverticulosis was shown to reduce both clinical pain and experimentally measured somatic hyperalgesia in patients with comorbid fibromyalgia [13,14]. And recently, the less invasive pain reprocessing therapy showed promise in decreasing auditory hyperresponsivity and modulating cortical activity in patients with chronic back pain [52]. Together with our earlier findings that experimental MMH is a predictor of postmenarcheal widespread pain [50], these results raise the possibility that MMH may be a useful marker for identifying which adolescents are most likely to benefit from early pelvic pain intervention. Prospective trials measuring MMH before and after pelvic pain intervention in adolescents are needed to test this directly.

Our sample, though representative of the surrounding communities, was skewed toward higher socioeconomic status and was generally healthy and pain-free, so the pelvic pain–MMH association may differ in more clinically or socioeconomically diverse populations. The sample was also entirely female. Rather than an incidental limitation, this reflects a deliberate design choice: somatic complaint and MMH are consistently higher in girls than boys around puberty [20,68], yet prior studies have not accounted for menstrual pain exposure as a contributor. By following an all-female cohort across the menarcheal transition with menstrual pain explicitly measured, we isolate this confound. Still, because our data cannot establish whether the pelvic pain–MMH relationship is sex-specific, mixed-sex cohorts are needed to confirm generalizability. A further limitation concerns the generalizability of the PCA solution itself. Component solutions from samples below approximately 500 can be unstable, particularly for weights (as opposed to the general pattern of loadings) [24,44]. Our baseline PCA sample (*n*=214) is below this threshold, and our cohort was demographically homogeneous (see Table 1), which may itself constrain the diversity of clustering patterns observed. Independent replication in a demographically distinct sample is an important next step. Reassuringly, supplementary attrition-sensitivity analyses indicate that this component structure was not detectably an artifact (see Supplemental Material). Future research would be well served to evaluate the factor structure and weighting scheme in larger more diverse samples to ensure replicability and generalizability of the results observed herein.

## Conclusion

To our knowledge, this is the largest study evaluating experimental MMH changes across the menarcheal transition. Non-menstrual pelvic pain, not menstrual pain, was most strongly associated with MMH, and this association was stable rather than time-dependent. This is consistent with MMH as an early, trait-like marker of pain vulnerability that co-occurs with pelvic pain from before menarche. Early characterization of sensory sensitivity may identify those at risk for future pelvic pain, while screening for pelvic pain may flag those with elevated MMH; both warranting closer monitoring across pubertal development.

## Competing Interests

Frank F. Tu reports royalties from Wolters Kluwer and stock options from Maipl Therapeutics. The remaining authors report no additional competing interests.

## Ethics Statement

All study procedures adhered to ethical guidelines outlined in the Declaration of Helsinki and received approval from Endeavor Health’s Institutional Review Board.

## Data Availability Statement

All data and code will be made available on Zenodo upon publication.

## Data Availability

All data and code will be made available on Zenodo upon publication.

## Acknowledgements

The authors thank Drs. G.F. Gebhart and Lynn S. Walker for advice on study design and interpretation. We acknowledge our laboratory staff for their efforts in coordinating and collecting data for the EMPATHY study: Ellen Garrison, Dina Vavarutsos, Katharine Jabaay, Emily Gal, Katlyn Dillane, and Genevieve Roth. We thank all the adolescents and parents/guardians for their participation. This work was supported by the Eunice Kennedy Shriver National Institute of Child Health and Human Development (R01HD096332).

