## Supplementary Material for "Multimodal hypersensitivity across the menarcheal transition is associated with pelvic pain in adolescents: a longitudinal study"

*Supplemental Material*

Matthew J. Kmiecik[1], Kevin M. Hellman[1,2], Sarah E. Darnell[1], Greg E. Miller[3], and Frank F. Tu[1,2]

[1]Department of Obstetrics and Gynecology, Endeavor Health, Evanston, IL, USA
[2]Department of Obstetrics and Gynecology, The University of Chicago Pritzker School of Medicine, Chicago, IL, USA
[3]Department of Psychology, Northwestern University, Evanston, IL, USA

Last modified: 05 Sep 2026

### Figures

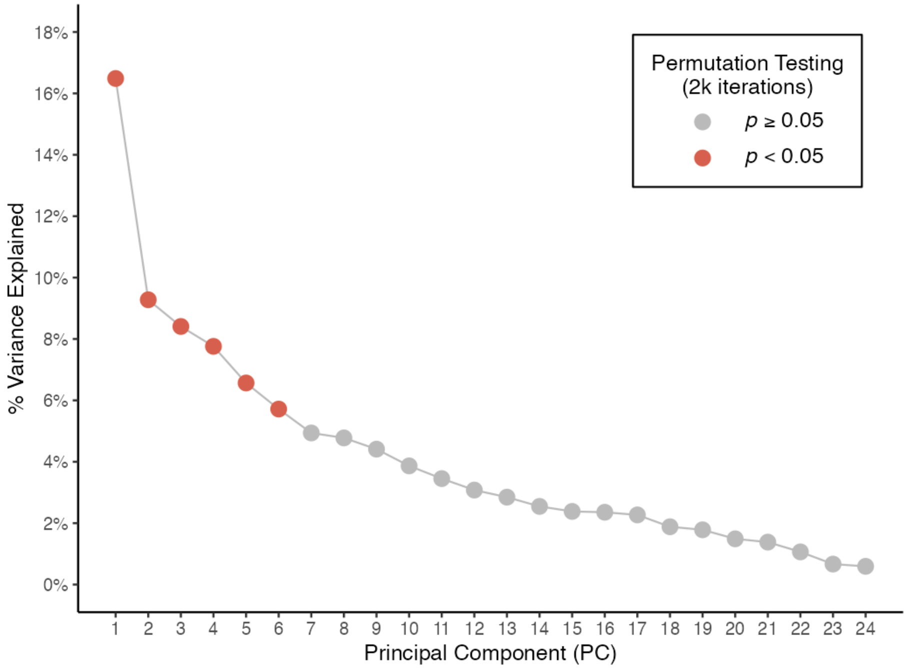

Figure S1: Scree plot indicating variance explained in each principal component and significance from permutations testing.

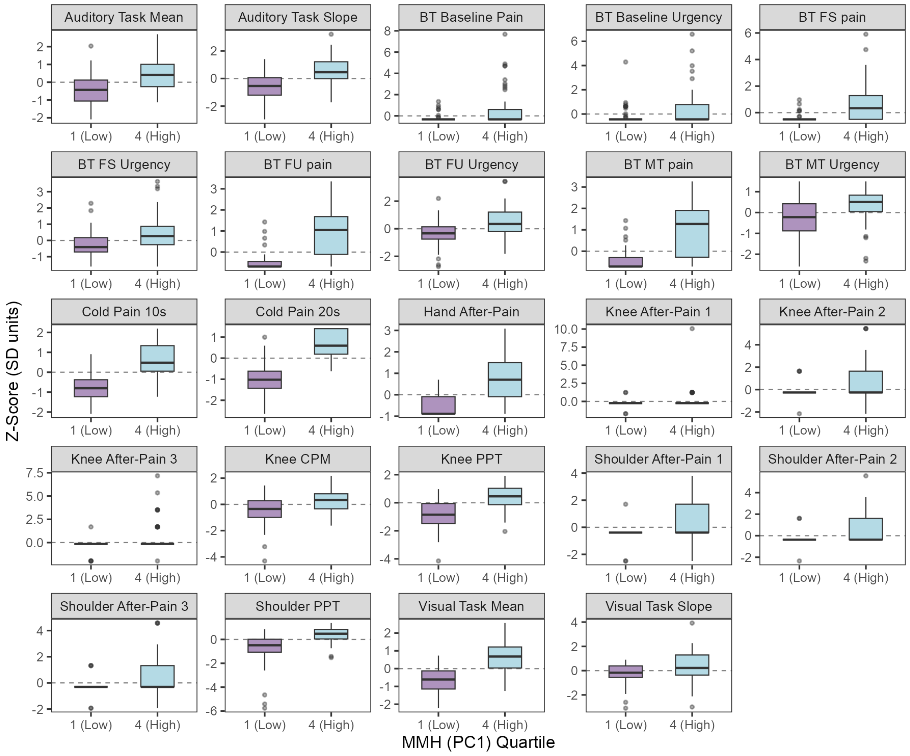

Figure S2: Boxplots of standardized multimodal hypersensitivity (MMH) testing measures. Participants were separated into low vs. high MMH based on quartiles of factor score loadings on the first principal component (PC1). Low MMH participants were in the bottom quartile and high MMH participants were in the upper quartile. Higher scores indicate greater sensitivity/impairment.

### Precision and Measurement Reliability

The main text reports no association between menstrual pain and multimodal hypersensitivity (MMH). This section reports the analyses behind the conclusion that this null is not attributable to insufficient sensitivity or to unreliable measurement of menstrual pain.

#### Bounding the menstrual pain association

The menstrual pain coefficient was $b$=0.070 PC1 units per NRS point, 95% CI [-0.039, 0.179] (Satterthwaite degrees of freedom). The upper confidence limit therefore bounds the association at 0.179 PC1 units per NRS point, or 28% of the pelvic pain estimate ($b$=0.644). Had menstrual pain been associated with MMH even one third as strongly as pelvic pain, the design would have detected it. Two one-sided tests (TOST) against a smallest effect of interest set at one

Table S1: Minimum detectable effect sizes at 80% power.

| Term | Observed $b$ | SE | MDES (raw) | MDES (SD) | Observed ≥ MDES |
|---|---|---|---|---|---|
| Time | -0.229 | 0.104 | 0.296 [0.291, 0.301] | 0.190 [0.187, 0.193] | No |
| Pelvic Pain | 0.644 | 0.126 | 0.362 [0.355, 0.368] | 0.229 [0.225, 0.233] | Yes |
| Menstrual Pain | 0.070 | 0.055 | 0.156 [0.153, 0.158] | 0.202 [0.199, 0.205] | No |

*Note.* MDES = minimum detectable effect size at 80% power, estimated as the 80% effective dose of a probit model fitted to simulated rejection rates across five effect sizes per term (800 simulations each; two-sided $\alpha$=.05; n=140 participants, 339 observations). Brackets give delta-method 95% confidence intervals. Raw MDES is in PC1 units per unit of the predictor (per year for time, per NRS point for menstrual and pelvic pain); standardized MDES rescales by SD(predictor)/SD(PC1). A raw MDES smaller than another term's indicates greater per-unit precision, not a larger effect.

third of the pelvic effect ($\Delta$=0.215) support that reading ($p$=0.005); against the more stringent $\Delta$=0.155 (0.2 *SD* of MMH per *SD* of menstrual pain) the equivalence test is marginal ($p$=0.063). Both $\Delta$ values are derived from the fitted model rather than fixed in advance, and both are reported so that the conclusion does not rest on a single choice of the smallest effect of interest.

### Detectable effect sizes

Holding the fitted model's variance components (between-subject intercept *SD*=1.22, random slope *SD*=0.29, residual *SD*=1.06) and the realized visit structure fixed, the fixed effect for each focal predictor was set in turn to each of five candidate values, and 800 datasets were simulated at every value, refitting the model and testing the focal term in each. Grids were centered on the analytic anchor $SE \times (t_{.975,df} + t_{.80,df})$ and spanned 0.7–1.3 times that value. The minimum detectable effect size (MDES) at 80% power was then estimated as the 80% effective dose of a probit model fitted to the simulated rejection rates across all five grid points, with a delta-method confidence interval.

Results are reported in Table S1. Per NRS point, the design was *more* sensitive to menstrual pain (MDES=0.156) than to pelvic pain (MDES=0.362), because menstrual pain was reported across the full 0–10 NRS range whereas pelvic pain was concentrated near zero. Any account of the null menstrual pain association that attributes it to lower sensitivity for that predictor is therefore unsupported. Placed on a common per-standard-deviation scale the two detection thresholds are close, marginally favoring menstrual pain (0.20 vs. 0.23 *SD*). These are the effects the design could detect, not the effects observed; the observed coefficients are compared directly below.

### Direct comparison of the two pain predictors

Per-unit coefficients for the two pain predictors are not directly comparable, because the predictors differ substantially in dispersion: menstrual pain was reported across the full 0–10 NRS range (subject-level *SD*=2.42) whereas pelvic pain was concentrated near zero (*SD*=1.18). The model was therefore refit with both pain predictors divided by their subject-level standard deviations, leaving the outcome in PC1 units, and the difference between the two per-*SD* coefficients was tested directly. The pelvic association was the larger of the two (difference=0.564 PC1 units per *SD* of predictor, *SE*=0.232, $t(123.4)=2.43$, $p=0.016$). The covariance between the two coefficients is negative, which widens rather than narrows the standard error of the contrast, so this test is conservative. Note that this contrast is not on the scale of the standardized $\beta$ column of Table 2 in the main text, which z-scores the outcome and time as well; dividing by *SD*(PC1)=1.863 converts it to that scale ($\beta=0.303$).

#### Test–retest reliability and attenuation

Menstrual pain was measured as a recall-based rating of average cramping pain over the preceding three months, so part of the null could in principle reflect measurement error rather than the absence of an association. Menstrual pain does not exist premenarche, so visits 1 and 2 are the only possible retest pair; among the 81 participants with a menstrual pain rating at both, test–retest reliability was moderate (ICC2=0.531 (0.356, 0.671); two-way random, absolute agreement, single measure). Classical disattenuation ($b$ / reliability) raises the menstrual coefficient from 0.070 to 0.132.

That correction over-corrects, and deliberately so. The retest interval here spans one to two years rather than a short reliability window, so the correlation conflates unreliability with genuine change in menstrual pain after menarche. The ICC is therefore a lower bound on reliability and the disattenuated coefficient an upper bound on the true association, not a point estimate of it. Read that way it strengthens rather than weakens the conclusion: even under the most generous correction these data permit, the pelvic association remains 4.9× larger than the menstrual one, i.e., the menstrual association is roughly one fifth the size of the pelvic one.

### Attrition Sensitivity Analyses

Only 119 of the 214 participants with a complete baseline multimodal hypersensitivity (MMH) battery contributed to the longitudinal mixed models reported in the main text, which drew on 140 participants in total. This section reports the analyses summarized briefly in the Methods and Results: how retained and non-retained participants compared at baseline, whether the MMH factor structure itself was invariant to retention, and whether the estimated developmental trajectory was distorted by attrition.

#### Retention taxonomy

The longitudinal mixed models drew on 140 participants. The mixed-model sample was not simply the baseline-PCA sample minus dropouts; however, 119 of the 140 mixed-model participants also had a baseline PCA score, while the remaining 21 did not complete the full baseline battery and therefore entered the mixed models through their follow-up visits alone. Restricting comparisons to the 214 baseline-PCA participants so that every participant considered has an observed baseline profile, 119 (55.6%) were retained in the mixed models, 58 (27.1%) never returned for a follow-up visit, 25 (11.7%) returned but never completed the MMH battery at a follow-up visit, and 12 (5.6%) returned with a complete follow-up MMH battery but were excluded from the published model because the visit-1 pain covariates of menstrual and pelvic pain were missing.

#### Baseline differences and their direction

Retained and non-retained participants (within the 214-participant baseline-PCA cohort) were compared on all 24 MMH battery measures, the four symptom questionnaires, and demographic/developmental measures (32 comparisons total; Welch's $t$-tests with Hedges' $g$, FDR-corrected within variable family), plus three categorical demographic comparisons ($\chi^2$ or Fisher's exact test). Only pubertal stage survived correction: retained participants had more advanced Tanner breast (2.63 vs. 2.28, $g$=0.44, $q$=.007) and pubic hair staging (2.55 vs. 2.22, $g$=0.37, $q$=.016) at baseline than non-retained participants. No MMH battery measure, questionnaire, or demographic variable (race, ethnicity, income) differed after correction (all $q \geq .11$).

Directly testing participants' baseline PC1 (MMH) factor scores told the same story, in the opposite direction from what a simple attrition account would predict: retained participants had somewhat *higher* baseline MMH than non-retained participants ($g$=0.33, 95% CI [0.06, 0.60], $q$=.016). In other words, participants who did not return for follow-up had lower, not higher, baseline MMH.

This difference was confined to PC1. A Hotelling's $T^2$ comparing the two groups across PC1–PC6 jointly was not significant ($T^2$=8.46, $F$(6,207)=1.38, $p$=.225, Mahalanobis $D$=0.40), nor was the corresponding test across all 24 components ($T^2$=17.92, $F$(24,189)=0.67, $p$=.880). PC1 was pre-specified as primary because it is the modeled outcome, so its per-component test does not require a significant omnibus to be interpreted; among the remaining permutation-significant components none approached a group difference (all $|g|$ 0.19, $q$=.771). An omnibus test spreads a single-component difference across the full set of dimensions, so its null result should be read as evidence that retention selected on MMH specifically rather than on the component structure broadly.

#### Factor-structure invariance

The analyses below compare principal-component solutions fit separately within the retained ($n$=119) and non-retained ($n$=95) baseline-PCA subgroups across the principal components (PCs) determined significant via permutation testing: PCs 1–6. In the non-retained subgroup, PC3–PC6 were separated by roughly one percentage point of explained variance each, so the individual component axes were only weakly identified by the data (i.e., rotationally indeterminate) even when the subspace they jointly span is well determined. We therefore applied an

Table S2: Permutation test of factor-structure invariance.

| Statistic | Observed | Null M | Null 2.5% | p | q |
|---|---|---|---|---|---|
| Tucker's $\phi$ (PC1) | 0.889 | 0.942 | 0.875 | 0.043 | 0.261 |
| Tucker's $\phi$ (PC2) | 0.763 | 0.763 | 0.513 | 0.437 | 0.812 |
| Tucker's $\phi$ (PC3) | 0.830 | 0.779 | 0.544 | 0.664 | 0.812 |
| Tucker's $\phi$ (PC4) | 0.693 | 0.768 | 0.482 | 0.212 | 0.636 |
| Tucker's $\phi$ (PC5) | 0.755 | 0.625 | 0.261 | 0.775 | 0.812 |
| Tucker's $\phi$ (PC6) | 0.663 | 0.509 | 0.188 | 0.812 | 0.812 |
| Subspace $R_V$ | 0.632 | 0.610 | 0.529 | 0.693 | |
| Correlation $R_V$ | 0.827 | 0.854 | 0.818 | 0.060 | |

*Note.* Retained (n=119) vs. non-retained (n=95) baseline-PCA participants, benchmarked against 2,000 random splits of the same 214 participants. Subspace $R_V$ is computed over PC1–PC6. Congruence coefficients are Procrustes-aligned. Null M and Null 2.5% are the mean and 2.5th percentile of the random-split null distribution. Lower values indicate less similar structure, so p is the proportion of random splits at least as dissimilar as the observed split. q is FDR-corrected across the six components.

orthogonal Procrustes rotation to remove this arbitrary axis orientation before computing Tucker's congruence coefficients ($\phi$) for the retained-vs-non-retained comparison.

Congruence coefficients have no absolute reference point at these sample sizes, so each observed statistic (the six component-wise $\phi$ values, the subspace $R_V$, and the correlation-matrix $R_V$) was benchmarked against a null distribution built from 2,000 random splits of the same 214 participants into groups of 119 and 95, matching the observed retained and non-retained group sizes, respectively (Table S2). After FDR correction, no statistic was less similar than chance splitting of the same data would produce, and for the more weakly identified components the observed congruence in fact exceeded the random-split average. PC1 was the one nominal exception ($\phi$=.889 vs. a random-split mean of .942, $p$=.044) and did not survive correction ($q$=.261); the correlation-matrix $R_V$ was likewise borderline ($p$=.060). Variance explained by PC1 was likewise nearly identical across the full sample, retained subgroup, and non-retained subgroup (16.5%, 17.5%, and 16.5%, respectively). Together, these results indicate that attrition had at most a marginal effect on the MMH factor structure, confined to PC1 and not surviving correction. The practical consequence is negligible: PC1 scores derived from a retained-only solution reproduce full-sample scores almost exactly ($r$=.994), so the outcome entering the mixed models is effectively unaffected.

**Robustness of the developmental trajectory to attrition**

Two complementary analyses tested whether this selection on baseline MMH level distorted the developmental trajectory reported in the main text. First, we extended the published mixed model with participation depth (dichotomized as contributing all three timepoints vs. fewer, since participants with a single timepoint carry no within-person slope information) and its interaction with time. Neither the main effect of depth ($F(1{,}129.8)=0.02$, $p=0.877$) nor its interaction with time ($F(1{,}114.7)=0.72$, $p=0.398$) was significant, indicating that the estimated rate of change in MMH did not depend on how much follow-up data a participant contributed.

Second, we predicted retention (yes/no) from participants' baseline PC1–PC6 scores and four questionnaires (logistic regression; all complete within the 214-participant cohort) and refit the published mixed model with stabilized inverse-probability-of-retention weights (IPW). No baseline predictor of retention survived FDR correction (all $q \geq .34$). Reweighting left the pelvic pain association essentially unchanged ($b=0.649$ unweighted vs. 0.647 IPW, $p<.001$ in both; Table S3). The time effect was more sensitive, and the sample restriction has to be separated from the weighting to read it correctly. Restricting the sample from the 140 participants of the published model to the 119 with a baseline principal component score moves the unweighted time effect from $p=.029$ (Table 2 of the main text) to $p=.043$; weighting within that subsample then moves it to $p=.061$ ($b=-0.229$ unweighted vs. $-0.211$ IPW). The weighted models are estimable only in this subsample, so the shift below conventional significance reflects its more limited power rather than a change introduced by the weights themselves. No coefficient shifted materially in magnitude (largest absolute change 0.026). The IPW-weighted model's random-effects covariance reached the boundary of the parameter space (intercept–slope correlation $= -1$); refitting with independent random intercepts and slopes shifted no fixed-effect estimate by more than 0.07, confirming the substantive conclusion does not depend on that degenerate covariance.

Table S3: Robustness of mixed-model coefficients to IPW weighting.

| Term | Published | Unweighted | p unw. | IPW | p IPW |
|---|---|---|---|---|---|
| Intercept | -0.225 | -0.270 | 0.069 | -0.296 | 0.048 |
| Tanner Breast (BP) | 0.028 | 0.088 | 0.699 | 0.108 | 0.635 |
| Tanner Breast (WP) | -0.002 | -0.045 | 0.746 | -0.026 | 0.846 |
| Tanner Hair (BP) | -0.240 | -0.355 | 0.116 | -0.371 | 0.101 |
| Tanner Hair (WP) | -0.042 | -0.011 | 0.931 | 0.002 | 0.986 |
| Time | -0.229 | -0.229 | 0.043 | -0.211 | 0.061 |
| Menstrual Pain | 0.070 | 0.054 | 0.364 | 0.050 | 0.408 |
| Pelvic Pain | 0.644 | 0.649 | $4.02 \times 10^{-6}$ | 0.647 | $5.71 \times 10^{-6}$ |
| Time × Menstrual Pain | 0.041 | 0.036 | 0.197 | 0.035 | 0.186 |
| Time × Pelvic Pain | -0.041 | -0.046 | 0.460 | -0.045 | 0.453 |
| Menstrual × Pelvic Pain | -0.022 | -0.016 | 0.755 | -0.020 | 0.705 |
| Time × Menstrual × Pelvic Pain | -0.008 | -0.004 | 0.852 | -0.009 | 0.684 |

*Note.* BP = between-person (grand-mean-centered); WP = within-person (visit-level deviation from the person mean). Menstrual and pelvic pain are grand-mean centered and time is centered at the mean interval of the visit at which pain was assessed, so lower-order coefficients are effects at sample-average pain and at that occasion; all three models use the centering constants from the published model so that estimates are directly comparable. Published estimates (b) are from the full mixed model on all 140 participants; Unweighted and IPW estimates are restricted to the 119 participants with a baseline principal component score. IPW estimates are from the original correlated-random-effects model, not the independent-random-slope sensitivity refit.
